# Health impact assessment of salt reduction in bread in Portugal: a pilot study

**DOI:** 10.1101/2021.12.20.21267610

**Authors:** Joana Santos, Joana Alves, Paula Braz, Roberto Brazão, Alexandra Costa, Mariana Santos, Ricardo Assunção, Teresa Caldas de Almeida, Luciana Costa

**Affiliations:** Department of Epidemiology, National Institute of Health Dr. Ricardo Jorge, Lisbon, Portugal; National School Public Health-Nova University of Lisbon, Lisbon, Portugal; NOVA National School of Public Health, Public Health Research Centre, NOVA University of Lisbon, Lisbon, Portugal; Comprehensive Health Research Center (CHRC). NOVA National School of Public Health, Universidade NOVA de Lisboa, Lisbon, Portugal; Food and Nutrition Department, National Institute of Health Dr. Ricardo Jorge, Lisbon, Portugal; Department of Health Promotion and Prevention of Non-Communicable Diseases, National Institute of Health Dr. Ricardo Jorge, Lisbon, Portugal; Portugal CESAM, Centre for Environmental and Marine Studies, University of Aveiro, Aveiro, Portugal; Faculty of Sciences, BioISI - Biosystems & Integrative Sciences Institute, University of Lisboa, Lisbon, Portugal

**Keywords:** HIA, Salt reduction program, Blood pressure, Hypertension

## Abstract

Hypertension is a risk factor for cardiovascular diseases, which can be caused by excessive salt intake. In Portugal, one of the main foods to contribute to ingestion of salt is bread. Thus, a voluntary Protocol was signed between stakeholders with the aim to reduce salt content in bread by 2021. Herein, a retrospective HIA was carried out to assess the impact in blood pressure (BP) after this agreement. In order to find average values of salt intake and BP in Portuguese population, national surveys were used. Also, estimates of BP reduction and its size effects were calculated based upon meta-analysis data. It is expected that salt intake will be reduced mostly in individuals with low educational level, men, aged between 65-74 years old and residents in South region of Portugal. Results in hypertensive patients indicate that a higher effect on BP will occur in the same profile of individuals, except age (between 55 and 64 years old). However, the estimated effect is very low for all groups, suggesting that the Protocol will contribute to modest health gains. Complementary measures supported by HIA studies need to be adopted to actively promote salt intake reduction and effectively prevent hypertension.

## 1. Introduction

Portugal is one of the European countries with the highest mortality rate due to stroke, while cardiovascular diseases (CVD) represent the main cause of mortality [1]. Hypertension (HT) was recognized as a major risk factor for the development of CVD, and is also one of the most relevant risk factor for reversing this condition [2–4]. On the other hand, the reduction of salt in food is one of the most important strategies to modify blood pressure (BP) and its impact on HT [5,6].

According to the National Food, Nutrition and Physical Activity Survey – IAN-AF (2015-2016), the Portuguese population has an average daily salt consumption of 7.4 g [7,8] despite the World Health Organization (WHO) recommends about 5 g of salt/day [9]. Also, bread and its by-products, for its high consumption, were considered one of the main foods to contribute to the ingestion of salt [8]. Thus, bread has been a subject of concern, especially due to the relationship between the quantities ingested and the development of HT, one of the greatest public health risks in Portugal [10].

In this respect, the national legislation has approached the strategy of reducing salt intake advocated by WHO and the European Commission. One of the principal approaches was to reduce salt content in bread. In fact, the Law nº. 75/2009 of 12th August established that the maximum salt content in bread after preparation would not exceed 1.4 g of salt per 100 g of bread. Subsequently, the Order 8272/2015 of 29th July established the creation of an Inter-Ministerial Working Group with the objective of proposing a set of measures for the reduction of salt consumption. Further, in November 2017, a collaborative agreement with the same purpose, hereinafter called ‘Protocol’, was signed between the General-Directorate of Health (DGS), the National Institute of Health Dr. Ricardo Jorge (INSA) and some Associations of Bakery, Pastry and Similar Commerce and Industries [11]. This coregulation agreement established new goals, phased and progressive, for the reduction of salt in bread, with the final goal of a maximum salt content of 1.0 g of salt per 100 g of bread in 2021. Also, it provided for a communication and awareness campaign to reduce salt consumption and promote the consumption of low salt bread at national level.

The objective of this work is to provide decision makers with evidence about the potential effects in health with the implementation of the ‘Protocol’, through a Health Impact Assessment (HIA) pilot study. In particular, we aimed to estimate the potential effect in BP from the implementation of this agreement to decrease the maximum salt content in bread in Portugal.

## 2. Materials and Methods

### 2.1. Study Design

A retrospective HIA with a focus on equity, in line with the goals agreed in the ‘Protocol’, was performed. This pilot study was coordinated by INSA under the supervision of WHO, following a “Learn by Doing” approach. The pilot was carried out during a “Capacity building HIA training Program” in the context of the Biennial Collaborative Agreement between the WHO and the Portuguese Ministry of Health.

### 2.2. HIA Procedure

In order to identify and predict health impacts associated with the ‘Protocol’, the classical HIA stages were considered in this work. A project team was created, including ten professionals from the major scientific domains related with the pilot study (Public Health, Nutrition, Education and Economy). This work group included representatives from the Departments of Health Promotion and Prevention of Noncommunicable Diseases, Food and Nutrition, and Epidemiology of INSA, as well as from the Regional Health Administration of Algarve, the Directorate-General for Education and NOVA National School of Public Health.

The HIA Guide developed by the Institute of Public Health in Ireland (IPH) [12] was used as documental reference. In particular, the *Screening* and *Scoping* tools were followed as suggested and are detailed below.

#### 2.2.1 Screening

In order to assess whether the ‘Protocol’ would have impact on health and establish the need to pursue the HIA, the *Screening tool* developed by IPH was used [12]. In this context, the HIA project team was invited to join a face-to-face meeting to discuss the various topics suggested and to meet a final consensus in this matter.

#### 2.2.2. Scoping

To set the scope of the HIA, a Steering Group (SG) was formed and the Terms of Reference (ToR) were established. The SG was composed by an advisory group of stakeholders to provide guidance, scientific and technical support. Stakeholders from salt and bread industry (Salexpor, Northern Bakery, Pastry and Similar Industry Association, Portuguese Distribution Business Association, Federation of the Portuguese Agri-Food Industry), from health (DGS -National Programme for Cardiovascular Diseases, INSA) and from other sectors of society as education (Directorate-General for Education) and consumers (Portuguese Association for Consumer Protection) were invited to be part of the SG and were requested to contribute with inputs and their views about the process. Additionally, a ‘Decision-Making Protocol’ was elaborated to frame the decision-making process under the SG, within the framework of the project. During this stage, selection and prioritization of the impacts to be assessed were also defined throughout the design of a causal chain.

#### 2.2.3. Appraisal

Data from two national surveys (National Food, Nutrition and Physical Activity Survey (IAN-AF) and the National Health Examination Survey (INSEF)) [8,13] were used to provide different but yet complementary information, necessary to achieve objectives of this study. Both surveys were conducted in 2015 in a representative sample of the Portuguese population. Further details on the questionnaire, sampling and fieldwork procedures are available on the projects methodological documents. IAN-AF ethical approval was obtained from the National Commission for Data Protection, the Ethical Committee of the Institute of Public Health of University of Porto and from the Ethical Commissions of each one of the Regional Administrators of Health. All participants were asked to provide their written informed consent for participation, according to the Ethical Principles for Medical Research involving human subjects expressed in the Declaration of Helsinki and national legislation. Written agreements from the parents were required for children and adolescents below 18 years old. For INSEF, the protocol was approved by the Ethics Committee of INSA and by the National Data Protection Authority (Authorization n. 9348/2010). All survey participants provided written informed consent.

The database from IAN-AF (n=3247) gathers individual data for daily salt consumption from bread while INSEF (n=4910) has individual information for BP measured by a health professional. Both have sociodemographic and health information regarding participants from 25 to 74 years old. To overcome the fact that it was not possible to link both surveys due to ethics and confidentiality, data was aggregated by sex, age group, region and educational level and then estimated the correspondent average salt consumption for each stratum. This also allowed us to estimate the average BP for each group and merge with IAN-AF data.

To estimate the expected daily salt consumption from bread we multiplied the average daily consumption for each stratum by 0.4 g that corresponds to the 29% salt content reduction expected with the ‘Protocol’ implementation from the current 1.4 g to 1.0 g of salt per 100 g of bread. Additionally, the average values of BP for each group were calculated using data from INSEF and were applied estimates from a meta-analysis showing that, for a decrease of 4.4 g in total salt intake, it is expected a decrease in BP of 2.4 mmHg for normotensive and 5.4 mmHg for hypertensive individuals [14]. Thus, the following formulas were applied:

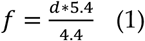

for hypertensive and

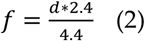

for normotensive individuals whereas (*f*) is the BP reduction expected after the implementation of the ‘Protocol’ and (*d*) is the correspondent absolute difference in salt intake.

Subsequently, a unique database was produced with current values of salt intake from bread, BP and its corresponding expected estimates after the implementation of the ‘Protocol’ measures to reduce salt content in bread. Estimates of the expected BP (*g*) were calculated by subtracting from the current systolic BP (*e*) the reduction estimated previously (*f*), according to the formula *g* = *e* − *f*. This was done both for hypertensive and normotensive individuals and according to the socio-demographic variables under analysis. Finally, the size effect was determined by dividing the expected reduction in BP (*f*) by the standard deviation of that group (*h*), according to the following formula:

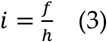

The statistical software R was used.

## 3. Results

### 3.1. Screening

After the *Screening* phase it was pointed out the importance of the ‘Protocol’ in relation to the potential health gains for the Portuguese population, since it could have a direct impact on the reduction of BP. Thus, the working group considered that health impacts were likely to occur and agreed to proceed with HIA.

### 3.2. Scoping

During the *Scoping* phase, the key health areas and populations likely to be the most affected by the policy measures set by the ‘Protocol’ under study were identified. Regarding the socioeconomic conditions, the working group considered that education, employment and community were the determinants of health most likely affected. In fact, previously collected data showed that individuals without any or low level of education, unemployed, retired and housewives are the ones with the highest prevalence of HT [15,16). Furthermore, males, elderly and population of Alentejo region are expected to be the most vulnerable groups and those who could have more beneficial effects on their health status because these are the groups that eat more bread. It was further considered the possible negative effects for the industry and trade linked to salt and bread sales, however these effects were devalued by the SG members. On the other hand, it was agreed that community interaction could be altered with the implementation of the ‘Protocol’. The awareness raising campaigns, aiming reducing the salt consumption and promoting bread consumption, could have considerable effects in changing habits of the population [17]. Moreover, qualitative changes in bread, as consequence of salt reduction measures, could have side effects in consumption pattern [18,19].

Additionally, in order to select the impacts to assess during the *Appraisal* phase, a causal chain was drawn and a pathway between bread consumption and potential health and economic effects was proposed (Figure 1). Following, the working group agreed to focus exclusively on the impact of salt reduction on BP, in an equity perspective. This decision was taken mainly due to time, human and financial resources constraints.

**Figure 1.**
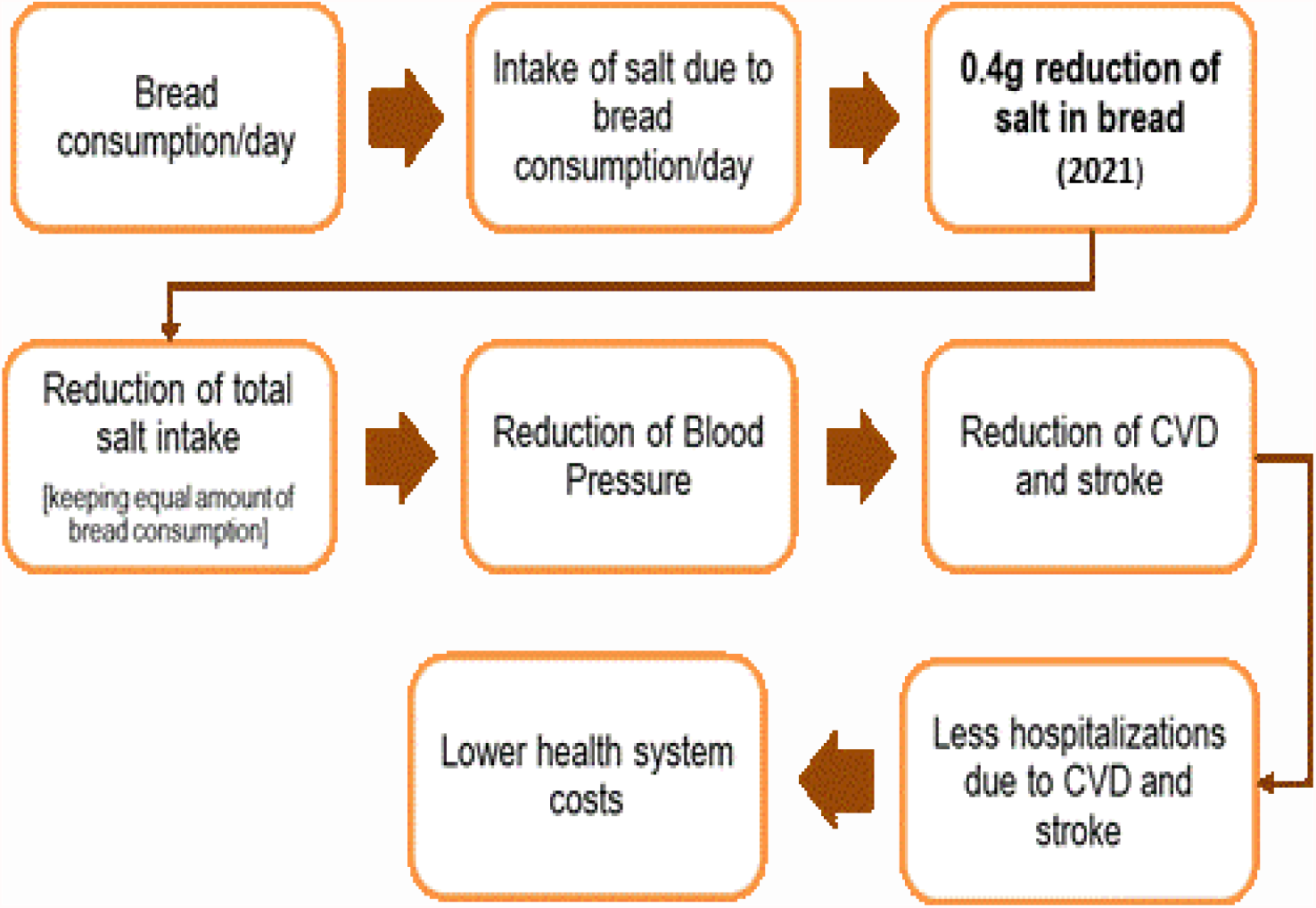
Causal chain drawn as the result of the scoping process.

### 3.3. Appraisal

#### 3.3.1. Effect in total salt intake

The decrease of 29% of the salt content in bread expected with the implementation of the ‘Protocol’, might reduce salt intake from 6.26 g to 5.94 g in women compared to a decrease from 8.88 g to 8.37 g in men, which would account for a reduction of 0.32 g and 0.51 g on total daily salt intake, respectively (Table I). Considering age group, the potential largest daily salt intake reduction is observed in individuals aged between 55 and 64 years old (7.44 g to 6.98 g) as well as in the individuals aged between 65 and 74 years old (6.75 g to 6.32 g). Considering educational level, it is estimated that the largest reduction in daily salt intake will occur in individuals with lower education. Individuals with primary education are expected to reduce total daily salt intake in 0.44 g (from 7.06 g to 6.62 g), while individuals with secondary education are estimated to show a reduction of 0.41 g (from 7.83 g to 7.42 g). Less impact of the ‘Protocol’ measures is expected for individuals with a university degree (reduction from 7.56 g to 7.20 g). Additionally, it is in the Alentejo region that the largest reduction in total daily salt intake is expected, from 8.55 g to 7.95 g, corresponding to a decrease of 0.60 g on total daily salt intake. In contrast, in Madeira region it is expected a lower reduction (0.30 g) corresponding to a decrease from 5.58 g to 5.28 g in total salt intake. This is consistent with the different daily salt intake patterns between the two regions, since the baseline salt intake was the lowest in Madeira and the highest in Alentejo.

**Table 1.** Expected mean reduction in salt intake after a 29% salt reduction in bread according to sex, age group, region and educational level.

|  | Salt in bread<br>(g/day-mean) | Current total<br>salt intake<br>(g/day-mean) | Expected total salt intake after<br>29% salt reduction in bread<br>(g/day-mean) | Absolute difference in salt intake<br>(g/day-mean) | Relative difference in salt intake (%) |
| --- | --- | --- | --- | --- | --- |
| **Sex** |  |  |  |  |  |
| Women | 1.11 | 6.26 | 5.94 | 0.32 | 5.1 |
| Men | 1.75 | 8.88 | 8.37 | 0.51 | 5.7 |
| **Age group** |  |  |  |  |  |
| 25-34 | 1.21 | 7.77 | 7.42 | 0.35 | 4.5 |
| 35-44 | 1.35 | 7.84 | 7.45 | 0.39 | 5.0 |
| 45-54 | 1.42 | 7.47 | 7.06 | 0.41 | 5.5 |
| 55-64 | 1.60 | 7.44 | 6.98 | 0.46 | 6.2 |
| 65-74 | 1.51 | 6.75 | 6.32 | 0.44 | 6.5 |
| **Region** |  |  |  |  |  |
| North | 1.35 | 7.84 | 7.44 | 0.39 | 5.0 |
| Center | 1.41 | 7.78 | 7.37 | 0.41 | 5.3 |
| Lisbon and Tagus Valley (LVT) | 1.27 | 7.77 | 7.41 | 0.37 | 4.8 |
| Alentejo | 2.07 | 8.55 | 7.95 | 0.60 | 7.0 |
| Algarve | 1.51 | 7.58 | 7.14 | 0.44 | 5.8 |
| Madeira | 1.04 | 5.58 | 5.28 | 0.30 | 5.4 |
| Azores | 1.33 | 7.25 | 6.87 | 0.39 | 5.4 |
| **Education level** |  |  |  |  |  |
| Primary | 1.50 | 7.06 | 6.62 | 0.44 | 6.2 |
| Secondary | 1.43 | 7.83 | 7.42 | 0.41 | 5.2 |
| University | 1.25 | 7.56 | 7.20 | 0.36 | 4.8 |

#### 3.3.2 Effect in blood pressure

In hypertensive patients (Table 2), with a mean daily salt intake expected to decrease 0.32 g in women and 0.51 g in men, it is estimated that the decrease in systolic BP will be higher in men (0.6 mmHg) than women (0.4 mmHg). However, a low effect size is anticipated for both groups.

**Table 2.** Impact of salt reduction on systolic BP in hypertensive patients according to sex, age group, region and education.

|  | Salt in bread<br>(g/day-mean)<br>(a) | Current total salt intake<br>(g/day-mean)<br>(b) | Expected total salt intake<br>(g/day-mean)<br>(c) | Absolute difference in salt intake<br>(g/day-mean)<br>(d) | Current systolic BP<br>(mmHg)<br>(e) | Reduction in the Systolic BP<br>(f) | Expected Systolic BP<br>(mmHg)<br>(g) | Standard Deviation of the group<br>(h) | Effect size<br>(i) |
| --- | --- | --- | --- | --- | --- | --- | --- | --- | --- |
| **Sex** |  |  |  |  |  |  |  |  |  |
| Women | 1.11 | 6.26 | 5.94 | 0.32 | 152.9 | 0.4 | 152.5 | 13.92 | 0.028 |
| Men | 1.75 | 8.88 | 8.37 | 0.51 | 151.9 | 0.6 | 151.3 | 12.19 | 0.051 |
| **Age group** |  |  |  |  |  |  |  |  |  |
| 25-34 | 1.21 | 7.77 | 7.42 | 0.35 | 147.3 | 0.4 | 146.9 | 7.06 | 0.061 |
| 35-44 | 1.35 | 7.84 | 7.45 | 0.39 | 148.1 | 0.5 | 147.6 | 9.29 | 0.052 |
| 45-54 | 1.42 | 7.47 | 7.06 | 0.41 | 153.1 | 0.5 | 152.6 | 12.94 | 0.039 |
| 55-64 | 1.60 | 7.44 | 6.98 | 0.46 | 152.8 | 0.6 | 152.2 | 13.63 | 0.042 |
| 65-74 | 1.51 | 6.75 | 6.32 | 0.44 | 153.5 | 0.5 | 153.0 | 13.39 | 0.040 |
| **Region** |  |  |  |  |  |  |  |  |  |
| North | 1.35 | 7.84 | 7.44 | 0.39 | 149.9 | 0.5 | 149.4 | 10.20 | 0.047 |
| Center | 1.41 | 7.78 | 7.37 | 0.41 | 151.7 | 0.5 | 151.2 | 12.23 | 0.041 |
| LVT | 1.27 | 7.77 | 7.41 | 0.37 | 152.8 | 0.5 | 152.3 | 12.27 | 0.036 |
| Alentejo | 2.07 | 8.55 | 7.95 | 0.60 | 154.8 | 0.7 | 154.1 | 15.28 | 0.048 |
| Algarve | 1.51 | 7.58 | 7.14 | 0.44 | 149.7 | 0.5 | 149.2 | 9.72 | 0.055 |
| Madeira | 1.04 | 5.58 | 5.28 | 0.30 | 152.9 | 0.4 | 152.5 | 13.70 | 0.027 |
| Azores | 1.33 | 7.25 | 6.87 | 0.39 | 154.2 | 0.5 | 153.7 | 14.69 | 0.032 |
| **Education level** |  |  |  |  |  |  |  |  |  |
| Primary | 1.50 | 7.06 | 6.62 | 0.44 | 153.6 | 0.5 | 153.1 | 13.70 | 0.039 |
| Secondary | 1.43 | 7.83 | 7.42 | 0.41 | 150.0 | 0.5 | 149.5 | 10.86 | 0.047 |
| University | 1.25 | 7.56 | 7.20 | 0.36 | 149.1 | 0.4 | 148.7 | 10.47 | 0.042 |

Considering the other variables under study, BP reduction is expected to be higher among individuals aged 55-64 years old (0.6 mmHg), population of Alentejo region (0.7 mmHg) and with the primary school (0.5 mmHg). A very small effect size is also estimated among all groups.

On the other side, estimates of the potential systolic BP decrease are expected to be very low for normotensive individuals, with magnitudes not exceeding the 0.3 mmHg reduction (Supplementary data-S1). The higher BP decrease is expected to follow the pattern estimated for the hypertensive patients. Accordingly, men, individuals aged 55 to 64 years old and population from South Portugal (in this case, Alentejo and Algarve) are expected to be the most positively affected. Nevertheless, and noteworthy, the effect size is significantly smaller than the estimated for hypertensives patients, showing very low values for all groups under analysis.

## 4. Discussion

The ‘Protocol’ under study aims to reach a 29% reduction of salt in bread, which leads to an expected reduction of 0.51 g and 0.32 g in total salt daily intake for men and women, respectively. However, our data showed that these changes seem to have slight effect in BP reduction. This might result from the fact that only aggregated data was available, so the potential impact could be diluted. Still, one can expect a larger reduction in salt intake among the population with higher consumption of bread, namely men, individuals aged 55-74 years old, population from Alentejo region and those with lower educational attainment. Concomitantly, after the implementation of the ‘Protocol’, these are the groups expected to have a higher decrease in systolic BP values, for hypertensive patients.

On the other hand, our findings show a low effect size for the all groups analysed indicating that the expected BP decrease caused by the reduction of salt intake through bread is too small to have an important impact in decreasing risk of CVD, according to previous data [14,20]. However, Barton *et al*. also suggests that despite no clear threshold value and due to the involvement of other risk factors, lowering BP leads to wide health benefits [20]. In fact, our results show that specific vulnerable groups of the population (men, elderly, population from South Portugal region and less educated individuals) are the ones expected to have the higher BP reduction.

Along with the direct impact in hypertension and CVD prevention, it is also important to take into consideration the putative economic effects related with the measures proposed in the ‘Protocol’. Several studies analyzed the economic efficiency of interventions aimed to reduce sodium intake. Selmer *et al*. observed that a reduction of the systolic BP by 1 mmHg can have a cost of 2040 US$ per life year saved [21]. Even interventions targeting a more conservative reduction of 0.5 g/day were generally cost effective, when considering the WHO benchmarks (cost effectiveness ratios lower than 3 times the Gross Domestic Product per capita) [22]. Thus, moderate reductions are generally cost-effective and interventions which reduce more than 2 mmHg could be cost-saving [21].

Nevertheless, despite the important value of actions encouraging food reformulation in achieving health gains, evidence suggests that only residual changes have been made in Portugal concerning the targets initially outlined [23]. In 2019, subsequently to the signature of the ‘Protocol’, an additional broad co-regulation agreement for reformulation of processed foods was signed including the reduction of salt in a range of different food categories, namely bread [24]. However, the available data demonstrates that voluntary initiatives have not been sufficiently effective in achieving reformulation targets, including salt intake reduction [25]. Previous studies postulate that mandatory approaches generate greater health gains than voluntary agreements [20,26]. In this context, the use of regulatory instruments, food and fiscal policies should be considered to sustain a health promotion strategy in this area. In fact, while Portugal is making a sound progress by the implementation of multiple actions with regard to food and nutrition, developed within the National Program of Healthy Eating [24] and the Integrated Strategy for the Promotion of Healthy Eating (EIPAS) [27], it is still uncertain the efficacy of the efforts to achieve public health targets through voluntary agreements.

In order to design the appropriate policy framework and to inform adequately political decision-making, studies of the possible impacts of actions taken on the various health outcomes are crucial. Herein, the projection models used to estimate the possible effects of salt reduction in bread in BP of Portuguese population showed that the measures proposed by the ‘Protocol’ are not sufficient to attain the ultimate public health targets. Adequate design of food policies could significantly benefit from the systematic use of HIA to meet the objectives set in the agenda to promote health and prevent Non-Communicable Diseases.

In this context, HIA could be a key methodological tool to support significantly the alignment of national strategies with the priorities established internationally, namely by WHO and European Commission.

## 5. Conclusions

The decrease in salt content in bread, endorsed by the voluntary ‘Protocol’ under analysis in this pilot study, is expected to contribute to a slight decrease in BP values in the Portuguese population. However, the effect of the measures agreed are expected to have a higher effect in specific vulnerable groups, which could contribute to reduce health inequalities in the community. Importantly, additional positive effects on health could be anticipated if other complementary and integrated measures are adopted. To this purpose, it is essential that decision makers acknowledge further the negative impact of salt consumption on health and turn this voluntary agreement into a compulsory act for all operators in the sector, especially bakery industry, bakery and bread retailers, including pre-packaged bread, but safeguarding eating goods with protected designation of origin. Also, it is recommended that similar regulatory acts could be extended to food products that are highly consumed by the Portuguese population, namely meat and its products, soups and other processed foods. These resolutions should be complemented by measures that increase consumers’ food and nutritional literacy such as public campaigns and nutrition labeling. Nonetheless, the requirement for a tight articulation between government agencies, health entities and partners from all sectors is needed, in order to effectively modify the food supply chain and a healthy eating environment is created. HIA, recognized as a beneficial tool to support and enable evidence based decisions, concerning health in all policies and intersectoral collaboration, should be used consistently and routinely in Portugal in order to design most effective food policies that promote health and reduce inequalities.

## Data Availability

All data produced in the present study are available upon reasonable request to the authors

## Supplementary Materials

One additional Table is provided.

## Author Contributions

JS contributed to the design of study, performed data analysis and wrote the first draft of the manuscript; JA reviewed literature and contributed to the writing of the article; PB, RB and MS contributed to the design of study, performed data analysis and reviewed the article; AC contributed to the writing and review; RA contributed to the design of study, performed data analysis, reviewed the article and supported edition; TCA supervised project administration, contributed to the study design and final review; LC supervised project administration, contributed to the study design and the writing of the first draft of manuscript, edited, revised and did the final review of document. All authors have read and agreed to the published version of the manuscript.

## Institutional Review Board Statement

This study uses secondary data. However, for IAN-AF ethical approval was obtained from the National Commission for Data Protection, the Ethical Committee of the Institute of Public Health of University of Porto and from the Ethical Commissions of each one of the Regional Administrators of Health. All participants were asked to provide their written informed consent for participation according to the Ethical Principles for Medical Research involving human subjects expressed in the Declaration of Helsinki and national legislation. Written agreements from the parents were required for children and adolescents below 18 years old. For INSEF, the protocol was approved by the Ethics Committee of INSA and by the National Data Protection Authority (Authorization n. 9348/2010). All survey participants provided written informed consent.

## Informed Consent Statement

Not applicable.

## Data Availability Statement

Data is available upon request for INSEF at National Health Institute Dr Ricardo Jorge. For IANAF the database is open and accessed online: www.ianaf.pt

## Acknowledgments

We acknowledge the support of all HIA team (particularly to Ana Cristina Guerreiro and Isabel Lopes representatives respectively of Regional Health Administration of Algarve and Directorate General of Education) as well as all the institutions and persons who shared information and knowledge throughout this project, particularly the representatives of the salt and bread industry (SALEXPOR, FIPA, AIPAN and APED). Special acknowledgments to Julia Nowacki, Gabriel Gulis, Marco Martuzzi and Jo Jewell for their valuable contribution on the development of this pilot study. RA also thanks to FCT/MCTES for the financial support to CESAM (UIDP/50017/2020+UIDB/50017/2020) through national funds.

